# Cervical cancer stem-like cell transcriptome profiles predict response to chemoradiotherapy

**DOI:** 10.1101/2020.11.03.20223339

**Authors:** Luciana W. Zuccherato, Christina M. T. Machado, Wagner C. S. Magalhães, Patrícia R. Martins, Larissa S. Campos, Letícia C. Braga, Andrea Teixeira-Carvalho, Olindo A. Martins-Filho, Telma M. R. F. Franco, Sálua O. C. Paula, Israel Tojal de Silva, Rodrigo Drummond, Kenneth J. Gollob, Paulo Guilherme O. Salles

## Abstract

Cervical cancer (CC) represents a major global health issue, particularly impacting women from resource constrained regions worldwide. Treatment refractoriness to standard chemo-radiotherapy has identified cancer stem cells as critical coordinators behind the biological mechanisms of resistance, contributing to CC recurrence. In this work, we evaluated differential gene expression in cervical cancer stem-like cells (CCSC) as biomarkers related to intrinsic chemoradioresistance in CC. A total of 31 patients with locally advanced CC and referred to Mario Penna Institute (Belo Horizonte, Brazil) from August 2017 to May 2018 were recruited for the study. Fluorescence-activated cell sorting (FACS) was used to enrich CD34+/CD45-CCSC from tumor biopsies. Transcriptome was performed using ultra-low input RNA sequencing and differentially expressed genes (DEGs) using Log2 fold differences and adjusted p value < 0.05 were determined. A panel of biomarkers was selected using the rank-based AUC (Area Under the ROC Curve) and pAUC (partial AUC) measurements for diagnostic sensitivity and specificity. The analysis showed 1062 DEGs comparing between the Non-Responder (n=10) and Responder (n=21) groups to chemoradiotherapy. Overlapping of the 20 highest AUC and pAUC values revealed five transcripts potentially implicated in innate chemoresistance (*ILF2, SNX2, COPZ1*, AC016722.1 and AL360175.1). This study identifies DEG signatures that serve as potential biomarkers in CC prognosis and treatment outcome, as well as identifies potential alternative targets for cancer therapy.

## 1. INTRODUCTION

Cervical Cancer (CC) is the second most frequent cancer and the fourth leading cause of cancer deaths in women worldwide. The Global Cancer Observatory (GLOBOCAN) estimates the burden of CC incidence in 2018 reached almost 570,000 women and a mortality rate of 54.6 % (311,365 patients). Approximately 85% of cases occur in low- and middle-income countries (LMICs), and are predominantly diagnosed in advanced stages.^1^ Despite the scientific advances in primary and secondary prevention (vaccine, HPV screening and precancerous lesions treatment, respectively), CC ranks as the fourth most common cause of cancer incidence and mortality in women worldwide. Impressively, around 50% of patients with CC died as consequence of treatment failure and other cancer-related complications. This unsuccessful scenario is also reflected in South American patients.^2^ This statistic strengthens the importance of developing novel therapeutic approaches for CC and achieving a more personalized medical care.

The heterogenous cellular composition of tumors leads to extreme genetic and epigenetic diversity and thereby produces a plethora of biological factors that can lead to a poor prognosis and low survival rates.^3^ Cancer Stem Cells (CSCs) are a pivotal participant in these processes. In normal tissues, stem cells are generally defined by a controlled self-renewal feature, with the ability to produce both specialized and undifferentiated tissue-maintaining cells. Conversely, CSCs can display perturbed growth properties, leading to cancer initiation, chemoresistance, and metastasis.^4^ Tumor heterogeneity can therefore be supported by the CSC paradigm, where a subset of cells, organized into hierarchical structures based on differentiation capacity, drive malignancy and therapeutic refractoriness.^5^

Given the importance of CSCs in cancer pathogenicity, it is not unexpected that many studies have sought to uncover the molecular pathways related to stemness. As observed in normal stem cells, CSCs exhibit genetic markers and pathways typically associated with proliferation.^6^ Aberrant expression of transcription factors SOX2, NANOG, OCT3/4, c-Myc,^7^ and disruption of Wnt/β-catenin, Hedgehog (Hh), Notch and PI3K/AKT/mTOR signaling pathways are representative hallmarks that sustain stem cell phenotype in CSC and support therapy resistance.^8^ Alternatively, mutations encompassing the tumor suppressor genes TP53, PTEN, and INK4A-ARF locus have been implicated in stem cell DNA damage pathways and self-renewal deregulation.^6^

Given the importance of CSCs in the tumorigenic process of solid tumors, we aimed to study differential expressed genes (DEG) in CCSC-like cells from tumor biopsies taken before treatment began in a cohort of CC patients that responded or not to chemoradiotherapy. The analysis of DEGs from sorted CCSC between responder vs. non-responder patients points to a set of potential biomarkers for prediction of response to chemoradiotherapy, as well as offering new insights into the potential role of CCSCs in therapy failure.

## 2. MATERIALS AND METHODS

### 2.1 Patient recruitment and sample selection

A total of 31 patients with CC and referred to Mário Penna Institute (Belo Horizonte, Brazil) from August 2017 to May 2018 were recruited to the study. Inclusion criteria was histopathological diagnosis of Squamous cell carcinoma (SCC) or Adenocarcinoma, no previous history of cancer or immune diseases. Cervical biopsies (FIGO stages II and III) were collected after participation agreement and signed the consent form. All patients were further submitted to radiotherapy concomitantly with chemotherapy, and clinical data were collected from medical records. This study was approved by the Ethics Committee at Instituto Mario Penna, Belo Horizonte, MG, which is registered with the Brazilian national ethics authority (CONEP). The study was approved (approval number: 1.583.784) for performing the prospective study with each patient signing a written informed consent before the initiation of any procedure related to the study.

Patients were screened as Responders (R) or Non-Responders (NR) based on Gynecologic Oncology parameters of absence/persistence of cervical lesions 8 months after the chemoradiotherapy. Clinical examination (vaginal, pelvic, abdominal), laboratorial analysis (cytology, new biopsy) and imaging (Ultrasound, Computer Tomography and Magnetic Resonance Imaging, when available) were assigned up to 8 months after treatment. Patients were considered R when cervical lesions were undetected after chemoradiotherapy. Patients with persistence of cervical lesions after the treatment (partial response, tumor progression or stable disease) were classified as NR.

### 2.2 Fluorescence Activated-Cell Sorting

Fluorescence-activated cell sorting (FACS) was used to isolate enriched Cervical Cancer Stem Cells-like (CCSCs) from a complex mixture of tumor cells based on their light scatter and fluorescent staining profiles. CC tissue fragments (5mm) from the 31 patients were fragmented using Med Machine® according to manufacturer’s instructions (BD-Biosciences). Cell suspension containing CCSCs was frozen in 20% HES cryoprotective solution (100 mL anhydrous glucose 1.7 g/L; Na(+1) 140 mEq/L; Cl(−1) 98 mEq/L; K(+1) 5 mEq/L; Mg (+2) 3 mEq/L; Gluconate 23 mEq/L; Acetate 27 mEq/L) and stored in liquid nitrogen until use. Considering the scarcity of CCSCs, two monoclonal antibodies, cell surface markers CD45 (APCH7 Clone 2D1) and CD34 (PE Clone 563) were used in the cell suspension for further FACS selection.

Cell concentration was increased to 5 ×10^6^ cells/mL. CCSCs-enriched subpopulations were isolated using FACS Aria® flow-sorter (BD-Biosciences). Yield mode was performed at 45 psi with 85-μm nozzle at a frequency of ∼51 kHz. Two fluorescence channels were analyzed (APC-H7 and PE). Cells were distinguished from debris in the sample by distinct FSC values, since debris can be identified as particles with lowest FSC values. Sorting of CCSCs was performed using a gate containing the CD45-/CD34+ population to eliminate contamination with hematopoietic stem cells, thereby enriching for CCSC-like cells (**Supplementary Figure 1**). Cells were considered positive above 10^2^ for each parameter based on negative populations defined by their autofluorescence. The CCSCs were sorted into cytometry tubes containing 1 μL of Lysis Solution (Lysis buffer and RNAse inhibitor from Takara® Kit Smart Seq V4 RNA) for genomic library construction and sequencing.

### 2.3 cDNA synthesis

SMART-Seq v4 Ultra-low Input RNA Kit for Sequencing (Takara Bio USA, CA) was used to generate the full-length cDNA from the selected sorted cells following the manufacturer’s instructions. Reactions with positive (Control Total RNA, provided by SMART-Seq v4 Ultra-low Input RNA Kit) and negative controls were carried out for quality control. Successful cDNA synthesis and amplification were considered when an Agilent High Sensitivity DNA Chip run on the Agilent Bioanalyzer 2100 (Agilent, CA) showed an electropherogram with a distinct peak spanning from 400 bp to 10,000 bp, and cDNA concentration ≥ 0.3 ng/μl detected with Qubit™ dsDNA HS Assay Kit on a Qubit 3 Fluorometer (Thermo Fisher Scientific, MA). Purified cDNAs were stored at −20°C for further processing.

### 2.4 Sequencing libraries

Library preparation of suitable cDNAs were performed using Nextera® XT Library Prep Kit (Illumina, CA) with Nextera® XT Index Kit V2 Set A (Illumina, CA). Samples were normalized to 40 pg/μl for a total 200 pg input of amplified cDNA. The protocol was performed as described by the manufacturer. Libraries were purified with 0.6 x bead ratio using Agencourt AMPure beads XP (Beckman Coulter, IN) and eluted in 52.5 μl of elution buffer. Quality parameters as size (440 bp average) and concentration (1.03 ng/μl average) were measured using High Sensitivity D1000 ScreenTape and reagents run on 4200 TapeStation System (Agilent, CA) and Qubit™ dsDNA HS Assay Kit on a Qubit 3 Fluorometer (Thermo Fisher Scientific, MA), respectively. Good quality libraries were normalized to 1 nM. Thirteen samples were pooled to further perform 101 cycles of single read sequencing using a NextSeq® 500/550 High Output Kit v2 (150 cycles) and NextSeq® 550 sequencer (Illumina, CA). Sixty-two libraries with more than 20 million reads were considered for analysis.

### 2.5 RNA-seq data analysis

Sequencing quality control and adapters were analyzed in the FastQ files using FastQC version 0.11.9. Trimming of the adaptor content and overrepresented sequences was performed using Trimmomatic.^9^ Quality check using FastQC was also performed on the trimmed sequences. Reads from the fastq files were aligned to the human reference (build GRCH380 and annotation file Homo_sapiens.GRCh38.83.gtf) using the 2-Pass protocol of the STAR software.^10^ The resulting alignment file was compressed, indexed and name-sorted using the samtools (version 0.1.19-44428cd). The count table was generated using GeneCounts mode from STAR. All steps are summarized in the **Supplementary Figure 2**; command lines and Pearl scripts of the workflow are available upon request.

### 2.6 Differential Expressed Genes (DEG) analysis and biomarkers selection

The count files were imported to DESeq2^11^ to perform differential expression analysis and an False Discovery Rate (FDR) adjusted p-value (q-value < 0.05) was used as the cutoff for assigning a given gene as a DEG between the groups R and NR. Benjamin and Hochberg procedure, implemented in DESeq2, were used additionally for p-values adjusting. The panel of biomarkers were proposed on the basis of the following criteria: (i) 20 highest values of AUC (area under the curve), (ii) 20 highest values of pAUC (partial area under the ROC curve statistic - (pAUC - 0.10) ^12^ and lastly, (iii) the common loci between the two selection criteria. All statistics were calculated using the R package “genefilter”.

### 2.7 Statistical analysis

Survival outcomes and the hazard ratio for disease progression or death were assessed via Kaplan-Meier methods and compared using Log-rank tests, using GraphPad Prism 5.0 (GraphPad Software Inc., La Jolla, Ca, USA). Forest plots represent the hazard ratio analysis of gene expression, obtained using a univariate Cox regression model in R (version 3.6.3, Rcore Team 2020; https://www.R-project.org), within survival package (version 3.2-3, Therneau 2020; https://CRAN.R-project.org/package=survival). Genes were selected by pAUC analysis.

## RESULTS

### 3.1 Clinicopathological characteristics of the cohort

Samples from 31 women with an average age of 52.3±16.8 years-old (range 24-82) were submitted to NGS analysis. Approximately 96% of patients had SCC, only one patient had adenocarcinoma, stages FIGO II-B (48.3%) and III-B (48.3%). The mean tumor size was 6.8 cm. Most patients had bilateral parametrial (71%) and vaginal involvement (90%) and tumor lesions were moderately (42%) or poorly differentiated (42%). The overall response was evaluated 8 months after the end of treatment and 21 patients were classified as Responders (R) and 10 as Non-responders (NR). Clinical information from the patients and histopathological analysis are summarized in **Supplementary Table 1**. Kaplan-Meier curves highlighted the poorer survival outcomes of NR patients with 12% survival at 12 months, whereas the R group showed 79% survival (Hazard Ratio [HR]: 6.44 95% CI: 0.11– 3.26, P?<0.0001). (**Figure 1**).

**Figure 1.**
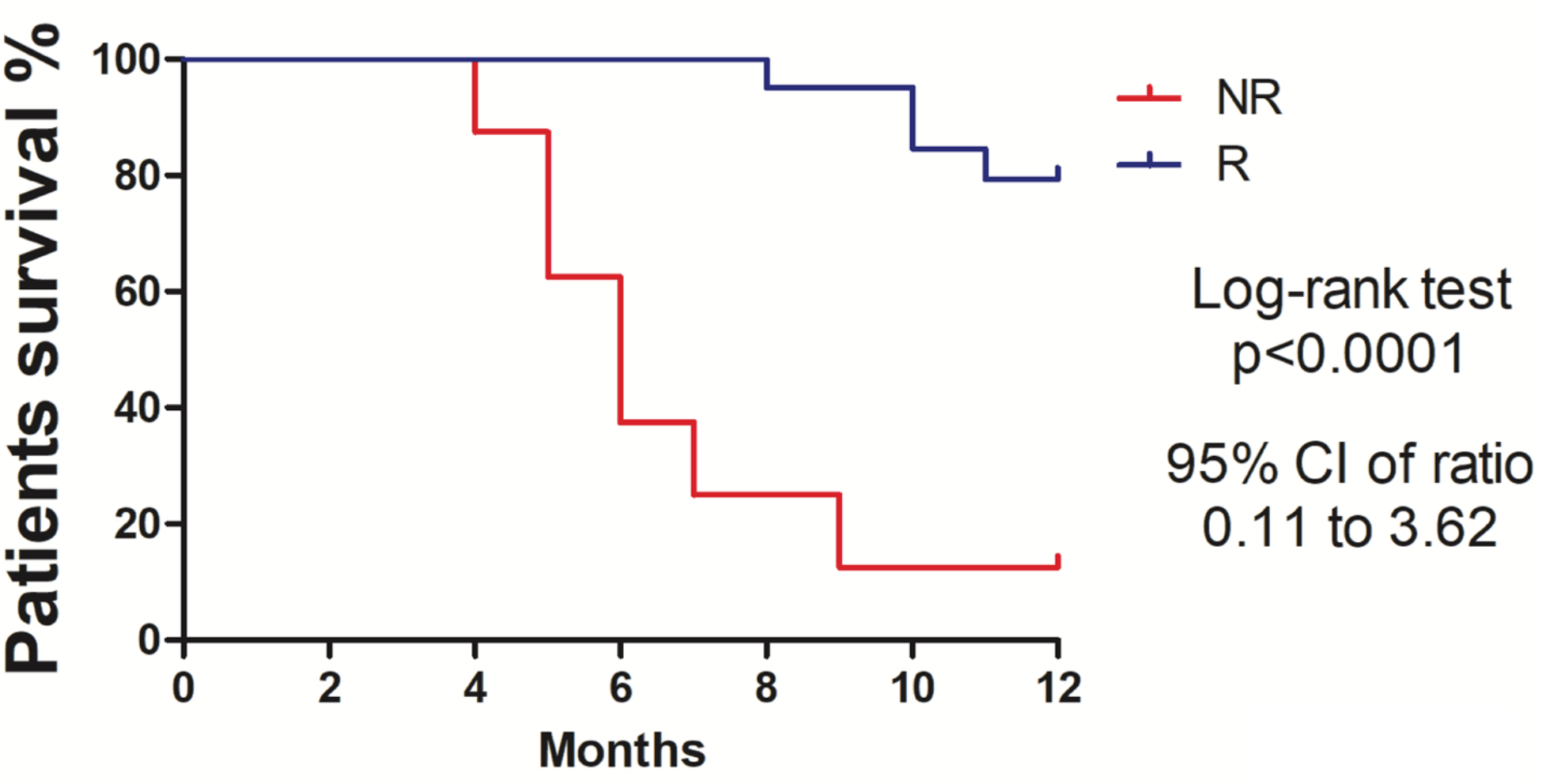
Kaplan-Meier survival curves showing higher percentage survival in Responder patients with cervical cancer. Log-rank test (Chi square 22.34, p<0.0001). Hazard ratio 0.64 (95%, CI 0.11-3.62).

### 3.3 Differential expressed genes (DEGs) in cervical cancer stem-like cells (CCSC)

DEGs between R and NR patients were analyzed using an adjusted p value of < 0.05. This analysis returned 1062 differentially expressed transcripts. Ninety-one percent (91%; 632/694) of the coding genes were underexpressed in NR patients; conversely, 85% of long non-coding RNAs (lncRNAs;125/147) and 82% of pseudogenes (128/155) were overexpressed in the NR group. Seventeen transcripts retrieved no annotations in the Ensembl Gene Set database, and therefore considered as novel in our analysis. The snoRNA (Small nucleolar RNA), Mt_tRNA (Mitochondrial transfer RNA), misc_RNA (miscellaneous RNA), miRNA (micro RNA), TEC (To be Experimentally Confirmed) and snRNA (Small nucleolar RNA) were grouped as “Other RNA” (n=49). The proportion of the DEGs and functional classes are detailed in **Figure 2A**.

**Figure 2.**
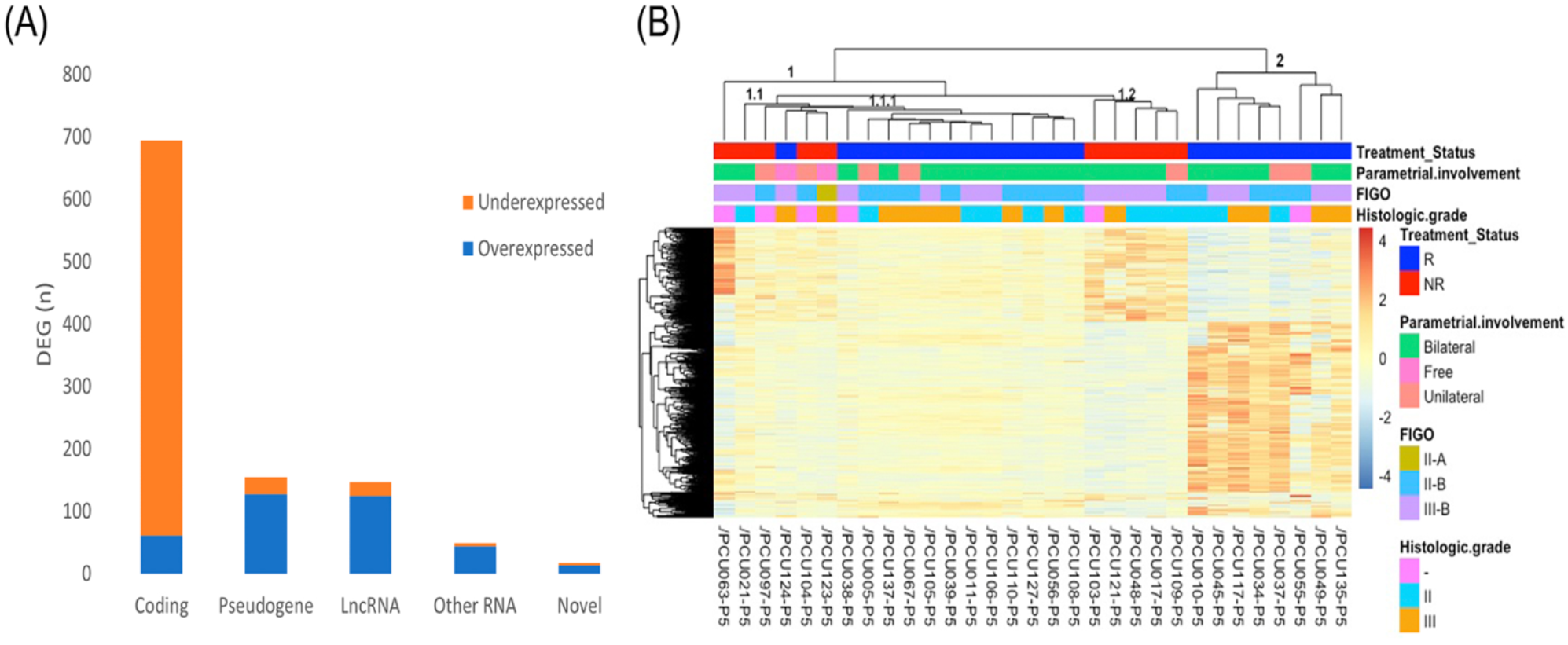
(A) Frequency of DEGs from CCSCs in Non-Responders (n=10) patients compared to Responders (n = 21). From the 1062 DEGs (adjusted p-value < 0.05), 694 protein coding, 155 pseudogenes, 147 long non-coding RNAs (LncRNA) and 49 transcripts classified as “Other RNA” (snoRNA, miRNA, miscRNA, TEC, snRNA and mitochondrial rRNA) were identified. Seventeen transcripts were classified as novel due to lack of reports in the Ensembl database (https://www.ensembl.org). Orange and blue colors represent the proportion of under and overexpressed DEG transcripts in each category, respectively. (B) Heatmap showing 1062 differential expressed genes (adjusted p-value < 0.05) in cervical cancer stem-like cells. Responder patients (R; n=21) are represented in blue and Non-Responders (NR; n=10) are colored in red. Clinical features of Parametrial involvement, FIGO Stages and Histological grades are detailed in the figure. The unsupervised clustering analysis of the DEGs revealed four clusters (1.1, 1.2, 1.1.1 and 2) corresponding to treatment status (R and NR).

An unsupervised clustering analyzes of the 1062 DEGs revealed four clusters related to the patient’s treatment outcome (NR vs. R) **(Figure 2B)**. A heterogeneous expression profile is apparent across the genes as exemplified by the heatmap. All patients segregated into clusters based on an expression pattern in agreement with the failure/success to chemoradiation treatment, with exception of one (PCU124, cluster 1.1). Clinicopathological characteristics of the patients, including pathologic parametrial involvement, FIGO stages and tumor pathological grades did not differentially segregate between the groups.

### 3.4. Long and small non-coding RNAs and epigenetic findings in CCSC

The long-noncoding RNAs (lncRNAs) differentially expressed in NR and R (n=147) represent 13.8% of all transcripts. From those, we detected 11 transcripts previously related to CC progression (**Table 1**) and 23 linked to diverse cancers (Supplementary Table 1). From the small non-coding transcripts (snoRNA, Mt_tRNA, TEC, misc_RNA, miRNA and snRNA), we identified two microRNAs (MIR765, MIR4779) and one snoRNA (SNORA12) with aberrant expression previously associated with cancer pathogenesis (**Table 1, Supplementary Table 2**).

**Table 1.** Long non-coding RNAs (lncRNA), small RNA (snoRNA**) and other genes differentially expressed in cervical cancer stem-like cells from Non-responders (NR) and Responders (R) related to cancer pathogenesis.

| Gene* | Log2FC | PADJ | Role in Cervical Cancer |
| --- | --- | --- | --- |
| *MIR205HG* | -3.86 | 0.0005 | Modulates biological activities of CC cells targeting SRSF1 and regulating KRT17.<sup>13</sup> |
| *MALAT1* | -3.52 | 0.0214 | High expression predicts a poor prognosis of CC,<sup>14</sup> promotes cancer cell growth and invasion/cisplatin resistance.<sup>15</sup> |
| *CRNDE* | -2.97 | 0.0060 | Promotes CC cell growth and metastasis.<sup>16</sup> |
| *SNHG8* | -2.86 | 0.0280 | Accelerates proliferation and inhibits apoptosis in HPV-induced CC.<sup>17</sup> |
| *NEAT1* | -2.64 | 0.0095 | Overexpression associated with radiosensitivity and CC poor prognosis.<sup>18</sup> |
| *SNHG6* | -2.46 | 0.0352 | Enhances the radio resistance and promotes growth of CC cells.<sup>19</sup> |
| *GAS5* | -2.08 | 0.0117 | Increases proliferation, migration<sup>20</sup>; low expression in the chemoradioresistant CC tissues.<sup>21</sup> |
| *NORAD* | -2.04 | 0.0143 | Significantly up regulated in CC tissues and cell lines.<sup>22</sup> |
| *LINC01133* | -1.31 | 0.0417 | Associated with CC progression.<sup>23</sup> |
| *LINC01048* | 2.44 | 0.0244 | High expression is an unfavorable prognostic factor for patients with squamous CC.<sup>24</sup> |
| *SNORA12\*\** | 3.32 | 0.0138 | Downregulated in CC.<sup>25</sup> |

\*Based on lncRNAs and small RNAs from public databases and literature, 11 transcripts associated with CC tumorigenesis were detected. Log2 fold change (Log2FC) values represent the difference in expression observed in NR when compared to R. (PADJ: Adjusted p-value <0.05; CC: cervical cancer).

### 3.4 Selection of Biomarkers

Based on the 1062 DEGs from the comparison between R and NR CCSC, we evaluated the prediction performance based on the partial area under the ROC curve statistic (pAUC; Pepe et al., 2013). The 20 transcripts with the highest pAUC (0.1) values and their cancer pathogenesis role are described in **Table 2**. Of these genes, three LcnRNA (*AL360091*.*3, AL360175*.*1* and *AC016722*.*1*); five pseudogenes (*RBM22P2, LOC100130121 (AC073324*.*1), OR1×1P, RPL7P52* and *MTND5P25*) and *BHMG1, SDS* and *METAZOA SRP* have not previously been shown to be transcripts in cancer.

**Table 2.** Description of the 20 highest values of pAUC (partial area under the ROC curve) from the differential expression genes of cervical cancer stem-like cells in patients with responsiveness (n=21) and failure (n=10) to chemoradioterapy (R *vs* NR).

| Gene | Gene name | Log2FG* | PADJ | GO Molecular function | Cancer pathogenesis |
| --- | --- | --- | --- | --- | --- |
| **ILF2\*\*** | Interleukin enhancer-binding factor 2 | -3.20 | 5.62E-09 | DNA/RNA binding | Overexpression associated with poor prognosis in CC.<sup>26</sup> |
| **EFNB1** | Ephrin-B1 | -2.04 | 0.0002 | Ephrin receptor binding | Induces invasion of ovary cancer.<sup>27</sup> |
| **FBXO34** | F-box only protein 34 | -1.90 | 0.0104 | Ubl conjugation pathway | - |
| **SNX2\*** | Sorting Nexin 2 | -1.86 | 0.0038 | Epidermal growth factor receptor binding | Silencing alters sensitivity to anticancer drugs targeted to c-Met and EGFR in lung cancer cells.<sup>28</sup> |
| **COPZ1\*\*** | Coatamer subunit zeta-1 | -1.82 | 0.0057 | Intra-Golgi vesicle-mediated transport | Knockdown causes Golgi apparatus collapse, blocked autophagy, induced apoptosis in proliferating and nondividing tumor cells.<sup>29</sup> |
| **CDC20** | Cell division cycle protein 20 homolog | -1.81 | 0.0051 | Ubiquitin ligase activator activity | Overexpression correlated with tumor grade and stage in CC.<sup>30</sup> |
| **DUSP16** | Dual specificity protein phosphatase 16 | -1.64 | 0.0221 | Phosphatase activity | Silencing causes cellular senescence by activating the tumor suppressors p53 and Rb.<sup>31</sup> |
| **OPA3** | Outer Mitochondrial Membrane Lipid Metabolism Regulator OPA3 | 1.07 | 0.0235 | Regulation of lipid metabolic process | High expression observed in pancreatic cancer tissues.<sup>32</sup> |
| **BHMG1 (AC074212.3)** | Meiosis Initiator | 1.36 | 0.0299 | DNA binding | - |
| **SDS** | Serine Dehydratase | 1.54 | 0.0394 | L-serine ammonia-lyase activity | - |
| **CCR4** | C-C chemokine receptor type 4 | 1.83 | 0.0234 | Chemokine binding | mRNA levels in local immune microenvironment of normal cervix was lower than in CC.<sup>33</sup> |
| **RBM22P2** | RNA Binding Motif Protein 22 Pseudogene 2 | 1.99 | 0.0099 | - | - |
| **AC073324.1 (RP11-339F13.2)** | Solute Carrier Family 19 Member 3 Pseudogene | 2.07 | 0.0493 | - | - |
| **AL360091.3 (RP11-384C4.7)** | LncRNA | 2.20 | 0.0143 | - | - |
| **AL360175.1 (RP11-524K22.1)\*\*** | LncRNA | 2.39 | 0.0020 | - | - |
| **OR1X1P** | Olfactory Receptor Family 1 Subfamily X Member 1 Pseudogene | 2.56 | 0.0033 | - | - |
| **METAZOA SRP** | Misc RNA | 2.77 | 0.0054 | - | - |
| **AC016722.1 (AC016722.3)\*\*** | LncRNA | 2.92 | 0.00089 | - | - |
| **RPL7P52** | Ribosomal Protein L7 Pseudogene 52 | 2.95 | 0.0005 | - | - |
| **MTND5P25** | MT-ND5 Pseudogene 25 | 3.13 | 0.0012 | - | - |

\*Log2 fold change (Log2FC) values represent the difference in expression observed in NR when compared to R. Transcripts are sorted by ascending number of Log2FC values. (PADJ: Adjusted p- value < 0.05; GO: Gene Ontology; CC: Cervical Cancer). \*\* The loci in common with AUC analysis.

The 20-top ranked pAUC genes from the CCSC DEGs separated into two groups using unsupervised cluster analysis, reflecting the treatment status between R and NR patients (**Figure 3A**). The gene expression profile of the patient PCU124 continued to cluster with the NR group as above mentioned with the 1062 DEGs.

**Figure 3.**
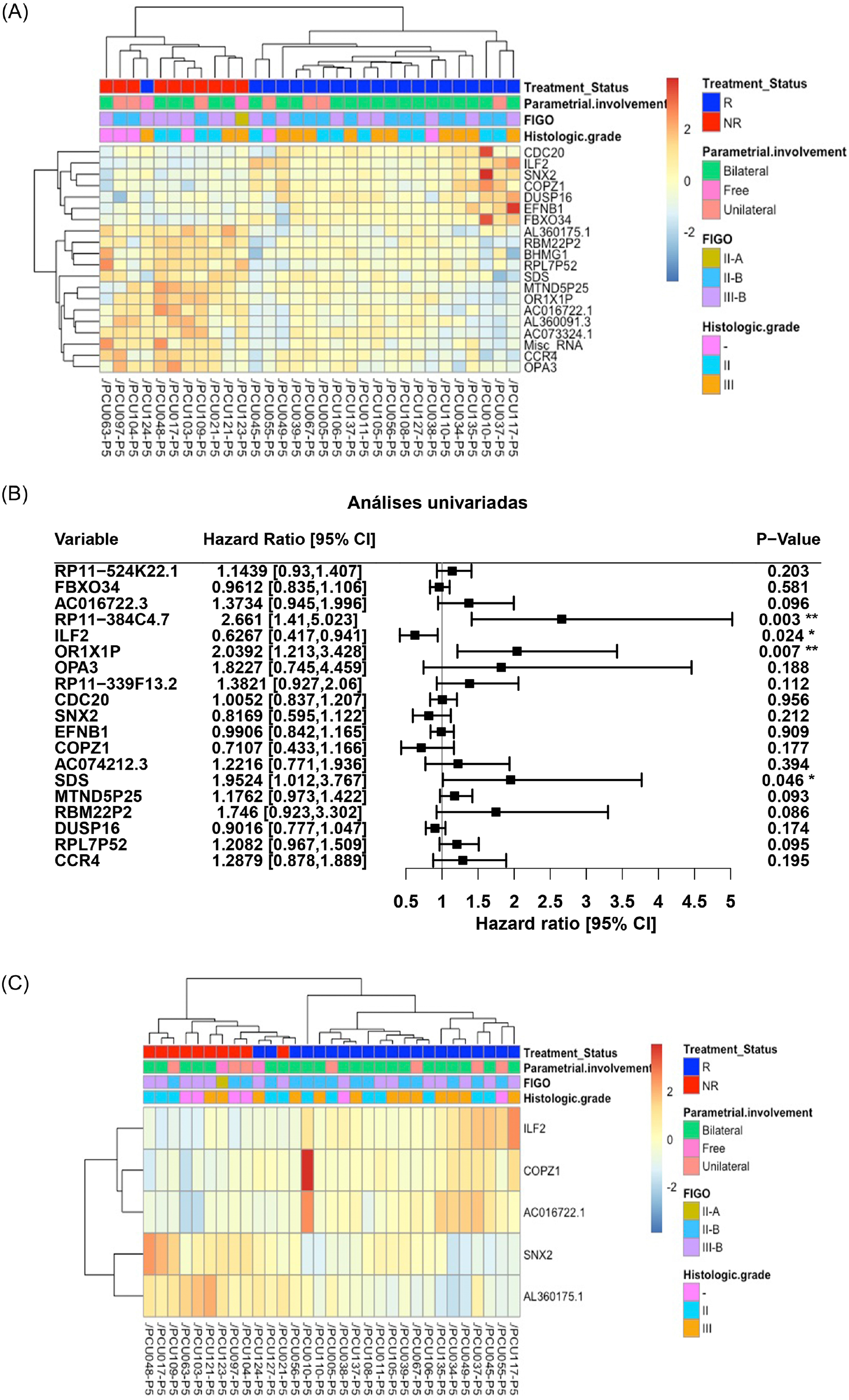
(A) Heatmap of the 20 genes with highest values of pAUC (0.1) from the 1062 differentially expressed transcripts (adjusted p-value < 0.05) in cervical cancer stem-like cells. Responders patients (R, n=21) are represented in blue and Non-Responders (NR; n=10) are represented in red. Clinical features of Parametrial involvement, FIGO stages and Histological grades are detailed. (B) Forest plot for prognostic effect of the 19 highest partial area under the ROC curve differential expressed genes in cervical cancer stem-like cells using a univariate analysis. (C) Heatmap showing the 5 overlapping differential expressed loci (adjusted p-value <0.05) between pAUC (0.1) and AUC in cervical cancer stem-like cells. Responders patients (R) are represented in blue and Non-Responders (NR) are represented in red. Clinical characteristics of parametrial involvement, FIGO stages and Histological grades are detailed in the figure.

The Hazard ratios (HR, 95% CI) for the 19 highest pAUC genes (excluding the Misc RNA) under a univariate analysis are shown in **Figure 3B**. Of those, the expression of *ILF2* showed a protection effect on patient survival (HR=0.63, p=0.024) while the genes *SDS* (HR=1.95, p=0.046) and *OR1×1P* (HR=2.04, p=0.007), and the lncRNA RP11-384C4.7 (HR=2.66, p=0.003) presented an increased risk with their increased expression.

To refine the number of robust biomarkers with potentially diagnostic utility, we also identified the highest 20 AUC values from the 1062 DEGs (**Supplementary Table 3**). Then, selecting the common genes between the pAUC and AUC analysis we identified 5 loci: *ILF2, SNX2, COPZ1*, ACO16722.1, AL360175.1 (**Figure 3C**). Unsupervised clustering of the selected DEGs maintained two clusters of genes, in agreement with NR and R status. The cluster enriched with R patients showed a higher expression of *ILF2, COPZ1* and AC016722.1, and lower values of Log2FC of *SNX2* and AL360175.1. The opposite pattern is seen for the NR group. Despite having a successful treatment outcome, PCU124,127, and 56 were grouped in the NR cluster.

To assess the quality parameters of prognostic prediction, accuracy, sensitivity, and specificity were evaluated using a univariate Logistic Regression (LR) for the 5 loci (**Table 3**). All parameters showed predictive values higher than 70%. The lncRNAs ACO16722.1 and AL360175.1 posed the highest accuracy (90.3%) and specificity (100% and 95.2%, respectively) evaluation; *ILF2* and AL360175.1 present the best sensitivity assessment (80%).

**Table 3.** Quality parameters of univariate Logistic Regression analysis. Five common loci between the highest 20 values of pAUC and AUC analysis from the differential expression genes in cervical cancer stem-like cells.

| Gene | Accuracy | Sensitivity | Specificity | Gene Function | Log2FC* |
| --- | --- | --- | --- | --- | --- |
| *ILF2* | 0.871 | 0.8000 | 0.9048 | Transcription factor for T-cell IL2 expression | -3.20 |
| AC016722.1 | 0.9032 | 0.7000 | 1.0000 | LncRNA | 2.91 |
| *SNX2* | 0.8387 | 0.7000 | 0.9048 | Intracellular protein trafficking | -1.85 |
| AL360175.1 | 0.9032 | 0.8000 | 0.9524 | LncRNA | 2.39 |
| *COPZ1* | 0.7419 | 0.5000 | 0.8571 | Autophagy and intracellular protein trafficking | -1.81 |

\*Log2 fold change values (Log2FC, adjusted p-value <0.05) represent the difference in expression in Non-responders (n=10) compared to Responders (n=21) to the chemoradiotherapy.

## 4. DISCUSSION

Nearly half a million new cases of CC occur each year, with the majority of cases being diagnosed in developing countries and at advanced disease stages.^1^ In addition, the number of deaths from CC is expected to increase to 410,000 by 2030. ^34^ It is evident that CC continues to be an important public health challenge worldwide. Despite screening programs and the recent advent of HPV vaccines, the quality of screening and treatment options must be improved. Based on cancer stem cell research and deep sequencing approaches, molecular alterations in CC have been widely explored including tumor heterogeneity, which is bringing new insights to clinical practice.

CSCs are self-renewing cancer cells responsible for expansion of the malignant mass in a dynamic process shaping the tumor microenvironment. CSCs have multiple differentiation capacities, independent anchorage growth, and chemoresistance.^35^ They represent approximately 0.1–10% of all tumor cells and express low levels of typical tumor-associated antigens compared to other tumor cells.^36^ After cytotoxic therapy regimens, residual tumor cells are enriched with CSCs, suggesting the importance of these cells in chemoresistance and disease relapse. CSCs may hijack the host immune surveillance to escape the toxic effects of chemotherapy and evasion of apoptosis, resulting in typically aggressive tumors with poor prognosis.^37^

CD34 is a sialomucin family-related transmembrane protein that is involved in the modulation of cell adhesion and signal transduction.^38^ While it was first identified as a hallmark of hematopoietic stem cells, studies demonstrated that CD34 is also present on nonhematopoietic cell lines, including embryonic fibroblasts and vascular endothelial progenitors. ^39^ Here we used CD34+ CD45-to sort subpopulations of tumor cells using flow cytometry as the most effective approach towards enriching stem-like tumor cells, thereby eliminating hematopoietic stem cells which are CD34+CD45+.

Based on the enriched CCSC from the bulk tumor biopsy, we conducted a low input RNA sequencing in patients with success (n=21) and refractoriness (n=10) to conventional chemoradiotherapy. The landscape of DEGs between the Non-responders (NR) and Responders (R) revealed 1062 loci (**Figure 2A**), characterized by coding, pseudogenes, long and small non-coding, and novel transcripts.

Interestingly, most of the coding DEGs of CCSC showed negative values of expression in NR in comparison to R (**Figure 2B**). The comprehensive pattern of underexpression in CCSCs might suggest a state of dormancy/quiescence, a key feature of tumor plasticity that protects stem-like cells from the antiproliferative agents.^40^. While CCSCs represent a key driving factor of tumorigenesis and metastasis, the majority maintain a quiescent or dormant state until changes occur in the microenvironment.^41^. Two main mechanisms account for tumor resistance to classical therapeutic approaches: Darwinian selection of cells harboring novel genetic variations (extrinsic resistance), or epigenetic events (chromatin remodeling and activation of pathways to cell stress), where dormancy and/or tumor quiescence can be achieved.^42, 43^

Indeed, epigenetic adaptations are deeply reflected in CCSCs resistance capabilities. Non-coding elements in the genome hold a diversity of regulatory factors responsible for the expression of proto-oncogenes or tumor-suppressor genes.^44^ Given that the deregulation of small and lncRNAs are strongly implicated in the tumorigenesis of CC, we performed a thorough investigation of these RNAs in our dataset. From the 24 DEGs reported (**Table 1; Supplementary Table 2**), the canonical oncogenic lncRNAs *MALAT1*,^14, 15^ *CRNDE*,^16^ *NEAT1*,^18^ and *NORAD* ^22^ showed negative expression values in NR as compared to R patients. Our finding differs from that seen in some of these studies, but it is important to remember that we were examining expression levels in purified stem-like cells, which represented a small percentage of cells in the entire tumor mass. Interestingly, decreased expression of the oncosupressive lncRNA *GAS5* is correlated with tumor development and worse clinical outcome in CC patients.^20,21^ These controversial findings are not unexpected in cancer studies, especially due to the molecular heterogeneity of tumors, sample source (cervical tissues, cell exfoliates, mucus, serum, cell cultures, and purified cell populations) and the variety of methodologies used. CSCs model and single-cell technologies provide an opportunity to study a heterogeneous collection of cells with distinct genetic and phenotypic properties within tumors and investigate what roles they play in these processes.^45^ The molecular mechanisms of non-coding RNAs in CC requires further characterization, particularly regarding the interplay between the diverse classes of RNAS and deregulation of metabolic pathways. Furthermore, the detection of these molecules in the serum of CC patients might lead to biosignatures of clinical relevance in non-invasive liquid biopsies.^46^

Here we focused on a set of loci with the highest potential for classifying responder vs. non-responder patients using a combination of pAUC and AUC analyses. The 20-loci signature selected using the pAUC analysis (**Table 2**) clearly separated the R and NR patients in a cluster analysis (**Figure 3A**). The transcriptional profile of the loci *ILF2, SDS, OR1×1P* and RP11-384C4.7 indicate significant prognostic capacity for CC pathogenesis (**Figure 3B**). The 5 loci that overlapped between the pAUC and AUC analysis led to a collection of loci that also segregated the R vs. NR groups: *ILF2, SNX2, COPZ1*, ACO16722.1, AL360175.1 (**Figure 3C**). The parameters of accuracy, specificity and sensitivity highlights the strong potential of these molecules to predict failure or success to chemoradiotherapy (**Table 3**). Single or multi-locus signature assays can be used to measure specific molecular pathway perturbations that could guide therapeutic decisions in the future. Thus, our approach using high-throughput RNAseq, where thousands of individual molecules were investigated, offers an initial set of biomarkers with the potential for clinical use upon further validation.

The Interleukin enhancer-binding factor 2 gene (*ILF2*, NF45) forms a complex with ILF3 (NF90) involved in transcription regulation,^47^ mitosis, ^48^ and DNA repair by nonhomologous end joining.^49^. *ILF2* acts as a tumor promoter, with overexpression associated with poor prognosis in CC, pancreatic ductal adenocarcinoma, non-small cell lung cancer and breast cancer. ^26 50^ Positive expression of *ILF2* may promote cancer occurrence and progression. However, our CCSC analysis revealed the opposite effect with *ILF2* overexpressed in CCSCs from patients with no CC recurrence after the chemoradiotherapy. This pattern of higher expression related to better survival in CC patients is also reported in The Human Protein Atlas database (p=0.087; https://www.proteinatlas.org/ENSG00000143621-ILF2/pathology/cervical+cancer).

In conclusion, we performed a comprehensive transcriptome analysis of CC stem-like cells enriched from fresh tumor biopsies. Despite numerous efforts, the discovery and establishment of new biomarkers for CC prognosis are lacking. Currently, overall prediction is mainly defined by clinical parameters. Thus, our results bring novel insights to the field. First, the landscape of intrinsic resistance to conventional chemoradiotherapy unveiled five distinguishing genes as novel putative biomarkers for predicting survival and response to therapy in CC patients. Second, we defined a distinct subset of non-coding transcripts in stem-like cells from CC, adding to our knowledge concerning the epigenetic factors driving treatment refractoriness. Third, the selected loci, in addition to standard clinical parameters, offer new insights towards prognostic assessment and therapeutic support in clinical practice. Importantly, further molecular characterization in a larger cohort of cervical cancer patients is required to validate our findings and possibly develop their use as clinically actionable biomarkers in the future.

## Data Availability

Datasets used in this study will be available upon reasonable request to the corresponding authors upon IRB review and approval for release of data to third parties.

## ACKNOWLEDGMENTS

We thank all the patients for participating in the study. We are also grateful to the Brazilian Funding Agencies of Ministry of Health (Pronon – Programa Nacional de Apoio à Atenção Oncológica; Grant number: NUP:25000.159953/2014-18) and the Minas Gerais State Research Agency (FAPEMIG; Grant number: TCT 19.011-13 - RT 00003-13). KJG is a CNPq fellow.

## Novelty and Impact

Cancer stem-like cells orchestrate tumor progression and strongly account for treatment resistance. We performed a comprehensive transcriptome analysis of sorted cervical cancer stem-like cells from fresh tumor biopsies. This analysis revealed five distinguishing genes as novel putative prognostic biomarkers in patients with cervical cancer.

## List of abbreviations (in alphabetical order)

CC: cervical cancer
CSC: cancer stem cell
CCSC: cervical cancer stem-like cells
CCNSC: Cancer non-stem cell
FACS: Fluorescence-activated cell sorting
DEG: Differentially expressed gene
Log2FC: Log2 fold change
AUC: Area under the curve
pAUC: partial area under the curve
ROC: Receiver operating characteristic curve
GLOBOCAN: Global Cancer Observatory
HPV: Human papillomavirus
RNA: Ribonucleotide
SCC: Squamous cell carcinoma
ADC: Adenocarcinoma
FIGO: International Federation of Gynecology and Obstetrics
FDR: False Discovery Rate
NR: Non-Responder
R: Responder
LR: Logistic Regression

## AUTHOR CONTRIBUTIONS

LWZ performed the NGS, contributed with the experimental design and wrote the paper. CMTM performed the FACS experiments, PRM contributed with patient recruitment, clinicopathological and survival analysis. LSC contributed with patient recruitment and FACS experiments, WCSM contributed with the experimental design and together ITS and RD performed the bioinformatics analysis, LCB contributed with paper writing, ATC and OAMF participated in FACS experimental design and analysis. TMRFF and SOCP were responsible for patient recruitment and clinicopathological data acquisition. KJG was responsible for study design, data analysis and paper writing, PGOS performed the anatomopathological analysis and contributed to experimental design and writing.

## SUPPLEMENTARY FILES

**Supplementary Figure 1.**
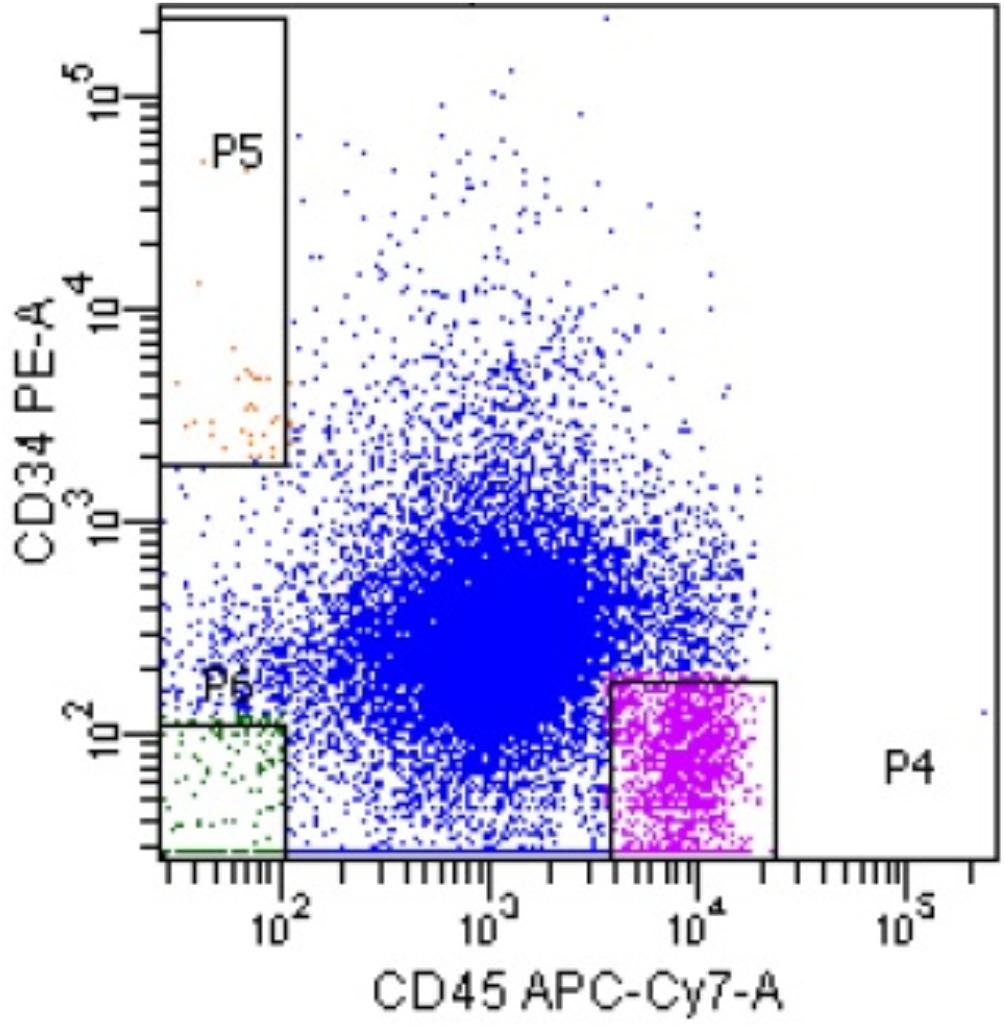
Fluorescence Activated-Cell Sorting representing the cellular profile and subpopulations of cervical cancer cell suspension. Cells were stained with anti-CD45-APC-Cy7 and anti-CD34-PE antibodies and sorted using a FACS ARIA cell sorter. The CCSCs were sorted based on CD34+CD45-(Gate P5) to avoid contamination with hematopoietic stem cells. The average number of cells sorted per sample was 2,000. The other cell populations were saved for future analysis.

**Supplementary Figure 2.**
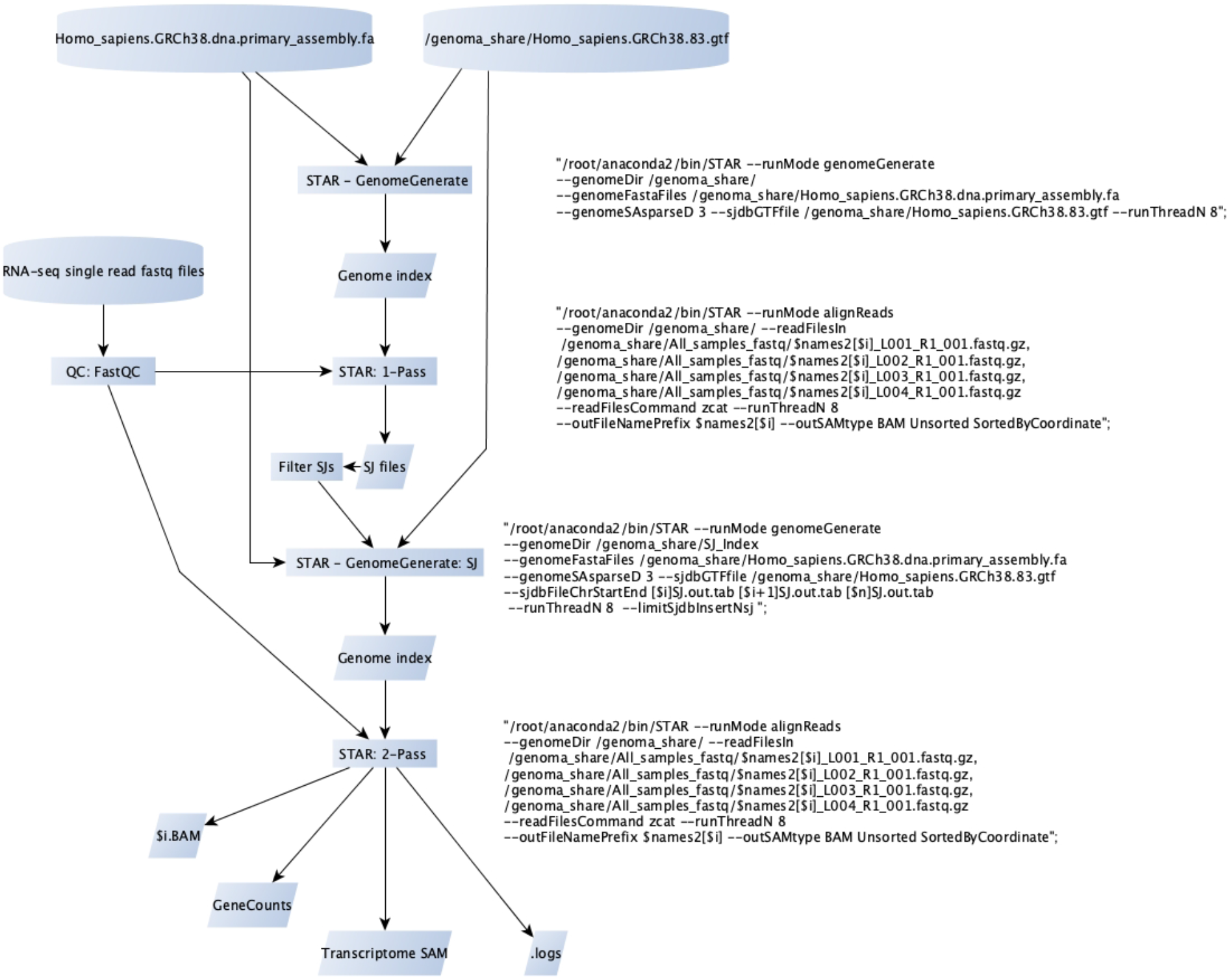
STAR alignment Workflow. The pipeline presents the different steps carried out in the pipeline, as well as the reference files (Human genome reference.fa and a .gtf file version) used during the process since fastq files until the matrix count (GeneCounts).

## Supplementary Tables

**Supplementary Table 1.** Clinicopathological characteristics and chemoradiotherapy response.

| Patient characteristics |  | N | Value* |
| --- | --- | --- | --- |
| Age | Years | 31 | 52.3±16.8 |
| Diagnostic | SCC | 30 | 96.8 |
|  | ICA | 1 | 3.2 |
| FIGO Stage | IIA | 1 | 3.2 |
|  | IIB | 15 | 48.4 |
|  | IIIB | 15 | 48.4 |
| Histological grade | II | 13 | 41.9 |
|  | III | 13 | 41.9 |
|  | NA | 5 | 16.1 |
| Parametrial involvement | Free | 1 | 3.2 |
|  | Unilaterally | 7 | 22.6 |
|  | Bilaterally | 22 | 71.0 |
|  | NA | 1 | 3.2 |
| Vaginal involvement | Yes | 28 | 90.3 |
|  | No | 1 | 3.2 |
|  | NA | 2 | 6.5 |
| Lymph node status | NX | 8 | 25.8 |
|  | N0 | 9 | 29.0 |
|  | N1 | 14 | 45.2 |
| Metastasis | MX | 10 | 32.3 |
|  | M0 | 9 | 29.0 |
|  | M1 | 12 | 38.7 |
| Tumor size (cm) | <4 | 1 | 3.2 |
|  | ≥4 | 29 | 93.5 |
|  | NA | 1 | 3.2 |
| Overall response | R | 21 | 67.7 |
|  | NR | 10 | 32.3 |

\*The values represent the mean ± standard error or %. SCC, Squamous Cell Carcinoma; ADC, Adenocarcinoma; NR, Non-Responders, R, Responders; NX, Regional lymph nodes cannot be assessed; N0, No regional lymph node metastasis; N1, Regional lymph node metastasis; MX, Distant metastasis cannot be assessed; M0, No distant metastasis; M1, Distant metastasis. NA, Not Available.

**Supplementary Table 2.** Long non-coding RNAs (lncRNA) and small RNA (miRNA*) differentially expressed in cervical cancer stem-like cells from Non-responders (NR) and Responders (R) related to cancer pathogenesis.

| Gene* | Log2FC** | PADJ | Role in cancer | Reference |
| --- | --- | --- | --- | --- |
| *PDCD4-AS1* | -2.91 | 0.0493 | Controls breast cancer progression by promoting tumor suppressor gene mRNA stability | 1 |
| *MAGI1-IT1* | -1.87 | 0.0357 | Promotes invasion and metastasis of epithelial ovarian cancer | 2 |
| *PRECSIT* | -1.69 | 0.0297 | Promotes progression of cutaneous squamous cell carcinoma | 3 |
| *FTX* | 1.07 | 0.0313 | Contributes to the progression of colorectal, gastric cancer and lung adenocarcinoma | 4-6 |
| *LINC00470* | 1.55 | 0.0320 | Promotes proliferation and invasion in glioma and hepatocellular carcinoma; attenuates chemosensitivity in glioma; oncogenic functions on gastric cancer cell | 7-9 |
| *LINC00974* | 1.71 | 0.0357 | Promotes cell cycle progression in gastric carcinoma and proliferation a metastasis in hepatocellular carcinoma | 10,11 |
| *LINC00449* | 1.71 | 0.0456 | Regulates the proliferation and invasion of acute monocytic leukemia | 12 |
| *MZF1-AS1* | 1.72 | 0.0270 | Induces the proline synthesis, tumorigenesis, and aggressiveness of neuroblastoma cells. | 13 |
| *MIR765\** | 2.01 | 0.0434 | Predicts a poor prognosis in patients with esophageal squamous cell carcinoma | 14,15 |
| *AC010789.1* | 2.13 | 0.0147 | Correlates with the prognosis of patients with colorectal cancer | 16 |
| *AC079341.1* | 2.20 | 0.0099 | Associated with the progression and prognosis of stage I hepatocellular cancer | 17 |
| *MIR4779\** | 2.34 | 0.0221 | Suppresses tumor growth by inducing apoptosis and cell cycle arrest | 18 |
| *AC012640.3* | 2.91 | 0.0006 | Expression can predict survival in hepatocellular carcinoma | 19 |

\*Based on lncRNAs and small RNAs from public databases and literature, 13 transcripts associated with cancer tumorigenesis were detected. \*\*Log2 fold change (Log2FC) values represent the difference in expression observed in NR when compared to R. PADJ: Adjusted p-value <0.05; CC: cervical cancer.

---

**Supplementary Table 3.** Description of the 20 highest values of AUC (area under the ROC curve) from the differential expression loci of cervical cancer stem-like cells in patients with responsiveness (Responder, n=21) and failure (Non-Responder, n=10) in the chemoradiotherapy.

| Transcript ID | Gene | Gene name | Log2FG* | PADJ |
| --- | --- | --- | --- | --- |
| ENSG00000251579 | **AC055733.3** | Pseudogene | -4.68 | 5.31E-04 |
| ENSG00000143621 | **ILF2\*** | Interleukin enhancer-binding factor 2 | -3.20 | 5.62E-09 |
| ENSG00000100028 | **SNRPD3** | Small Nuclear Ribonucleoprotein D3 Polypeptide | -3.14 | 9.06E-05 |
| ENSG00000251340 | **MTCYBP35** | Mitochondrially Encoded Cytochrome B Pseudogene 35 | -3.13 | 1.00E-03 |
| ENSG00000119396 | **RAB14** | RAB14, Member RAS Oncogene Family | -2.37 | 2.83E-03 |
| ENSG00000125871 | **MGME1** | Mitochondrial Genome Maintenance Exonuclease 1 | -2.35 | 4.34E-03 |
| ENSG00000177733 | **HNRNPA0** | Heterogeneous Nuclear Ribonucleoprotein A0 | -1.93 | 3.23E-03 |
| ENSG00000178974 | **FBXO34** | F-Box Protein 34 | -1.90 | 1.04E-02 |
| ENSG00000205302 | **SNX2\*** | Sorting Nexin 2 | -1.86 | 3.84E-03 |
| ENSG00000111481 | **COPZ1\*** | Coatomer subunit zeta-1 | -1.82 | 5.70E-03 |
| ENSG00000116786 | **PLEKHM2** | Pleckstrin Homology and RUN Domain Containing M2 | -1.44 | 8.69E-03 |
| ENSG00000251629 | **LINC02241** | LncRNA | 1.32 | 1.41E-02 |
| ENSG00000213468 | **FIRRE** | LncRNA | 1.40 | 3.33E-02 |
| ENSG00000178229 | **ZNF543** | Zinc Finger Protein 543 | 1.54 | 8.85E-03 |
| ENSG00000279632 | **AP003108.4** | TEC | 1.85 | 2.16E-02 |
| ENSG00000259747 | **AC102941.1**<br>**(RP11-275I4.2)** | LncRNA | 2.06 | 2.98E-03 |
| ENSG00000280422 | **AC115284.2** | TEC | 2.11 | 2.00E-02 |
| ENSG00000233470 | **AL360175.1\*** | LncRNA | 2.39 | 2.01E-03 |
| ENSG00000273791 | **AC007204.1** | BCL2/Adenovirus E1B 19kDa Interacting Protein 3 (BNIP3) Pseudogene | 2.67 | 4.70E-04 |
| ENSG00000226548 | **AC016722.1\*** | LncRNA | 2.92 | 8.85E-04 |

\*Log2 fold change (Log2FC) values represent the difference in expression observed in Non-responders compared to Responders. Transcripts are sorted by ascending number of Log2FC. PADJ: Adjusted p-value < 0.05; LncRNA: long non-coding RNA; TEC: To be Experimental Tested.

## REFERENCES

1. Ferlay J, Colombet M, Soerjomataram I, Mathers C, Parkin DM, Pineros M, Znaor A, Bray F. Estimating the global cancer incidence and mortality in 2018: GLOBOCAN sources and methods. Int J Cancer 2019;144: 1941–53.

2. Arbyn M, Weiderpass E, Bruni L, de Sanjose S, Saraiya M, Ferlay J, Bray F. Estimates of incidence and mortality of cervical cancer in 2018: a worldwide analysis. Lancet Glob Health 2020;8: e191–e203.

3. Gatenby RA, Brown JS. Integrating evolutionary dynamics into cancer therapy. Nat Rev Clin Oncol 2020;17: 675–86.

4. Valent P, Bonnet D, De Maria R, Lapidot T, Copland M, Melo JV, Chomienne C, Ishikawa F, Schuringa JJ, Stassi G, Huntly B, Herrmann H, et al. Cancer stem cell definitions and terminology: the devil is in the details. Nat Rev Cancer 2012;12: 767–75.

5. Rich JN. Cancer stem cells: understanding tumor hierarchy and heterogeneity. Medicine (Baltimore) 2016;95: S2–7.

6. Visvader JE, Lindeman GJ. Cancer stem cells in solid tumours: accumulating evidence and unresolved questions. Nat Rev Cancer 2008;8: 755–68.

7. Mendoza-Almanza G, Ortiz-Sanchez E, Rocha-Zavaleta L, Rivas-Santiago C, Esparza-Ibarra E, Olmos J. Cervical cancer stem cells and other leading factors associated with cervical cancer development. Oncol Lett 2019;18: 3423–32.

8. McCubrey JA, Rakus D, Gizak A, Steelman LS, Abrams SL, Lertpiriyapong K, Fitzgerald TL, Yang LV, Montalto G, Cervello M, Libra M, Nicoletti F, et al. Effects of mutations in Wnt/beta-catenin, hedgehog, Notch and PI3K pathways on GSK-3 activity-Diverse effects on cell growth, metabolism and cancer. Biochim Biophys Acta 2016;1863: 2942–76.

9. Bolger AM, Lohse M, Usadel B. Trimmomatic: a flexible trimmer for Illumina sequence data. Bioinformatics 2014;30: 2114–20.

10. Dobin A, Gingeras TR. Mapping RNA-seq Reads with STAR. Curr Protoc Bioinformatics 2015;51: 11 4 1–4 9.

11. Love MI, Huber W, Anders S. Moderated estimation of fold change and dispersion for RNA-seq data with DESeq2. Genome Biol 2014;15: 550.

12. Pepe MS, Kerr KF, Longton G, Wang Z. Testing for improvement in prediction model performance. Stat Med 2013;32: 1467–82.

13. Dong M, Dong Z, Zhu X, Zhang Y, Song L. Long non-coding RNA MIR205HG regulates KRT17 and tumor processes in cervical cancer via interaction with SRSF1. Exp Mol Pathol 2019;111: 104322.

14. Zhang Y, Wang T, Huang HQ, Li W, Cheng XL, Yang J. Human MALAT-1 long non-coding RNA is overexpressed in cervical cancer metastasis and promotes cell proliferation, invasion and migration. J BUON 2015;20: 1497–503.

15. Wang N, Hou MS, Zhan Y, Shen XB, Xue HY. MALAT1 promotes cisplatin resistance in cervical cancer by activating the PI3K/AKT pathway. Eur Rev Med Pharmacol Sci 2018;22: 7653–9.

16. Meng Y, Li Q, Li L, Ma R. The long non-coding RNA CRNDE promotes cervical cancer cell growth and metastasis. Biol Chem 2017;399: 93–100.

17. Qu X, Li Y, Wang L, Yuan N, Ma M, Chen Y. LncRNA SNHG8 accelerates proliferation and inhibits apoptosis in HPV-induced cervical cancer through recruiting EZH2 to epigenetically silence RECK expression. J Cell Biochem 2020;121: 4120–9.

18. Han D, Wang J, Cheng G. LncRNA NEAT1 enhances the radio-resistance of cervical cancer via miR-193b-3p/CCND1 axis. Oncotarget 2018;9: 2395–409.

19. Liu J, Liu X, Li R. LncRNA SNHG6 enhances the radioresistance and promotes the growth of cervical cancer cells by sponging miR-485-3p. Cancer Cell Int 2020;20: 424.

20. Yan Z, Ruoyu L, Xing L, Hua L, Jun Z, Yaqin P, Lu W, Aili T, Yuzi Z, Lin M, Huiping T. Long non-coding RNA GAS5 regulates the growth and metastasis of human cervical cancer cells via induction of apoptosis and cell cycle arrest. Arch Biochem Biophys 2020;684: 108320.

21. Fang X, Zhong G, Wang Y, Lin Z, Lin R, Yao T. Low GAS5 expression may predict poor survival and cisplatin resistance in cervical cancer. Cell Death Dis 2020;11: 531.

22. Huo H, Tian J, Wang R, Li Y, Qu C, Wang N. Long non-coding RNA NORAD upregulate SIP1 expression to promote cell proliferation and invasion in cervical cancer. Biomed Pharmacother 2018;106: 1454–60.

23. Ding S, Huang X, Zhu J, Xu B, Xu L, Gu D, Zhang W. ADH7, miR-3065 and LINC01133 are associated with cervical cancer progression in different age groups. Oncol Lett 2020;19: 2326–38.

24. Chen L, Chen Q, Kuang S, Zhao C, Yang L, Zhang Y, Zhu H, Yang R. USF1-induced upregulation of LINC01048 promotes cell proliferation and apoptosis in cutaneous squamous cell carcinoma by binding to TAF15 to transcriptionally activate YAP1. Cell Death Dis 2019;10: 296.

25. Roychowdhury A, Samadder S, Das P, Mazumder DI, Chatterjee A, Addya S, Mondal R, Roy A, Roychoudhury S, Panda CK. Deregulation of H19 is associated with cervical carcinoma. Genomics 2020;112: 961–70.

26. Shi C, Yang Y, Yu J, Meng F, Zhang T, Gao Y. The long noncoding RNA LINC00473, a target of microRNA 34a, promotes tumorigenesis by inhibiting ILF2 degradation in cervical cancer. Am J Cancer Res 2017;7: 2157–68.

27. Iwasaki K, Ninomiya R, Shin T, Nomura T, Kajiwara T, Hijiya N, Moriyama M, Mimata H, Hamada F. Chronic hypoxia-induced slug promotes invasive behavior of prostate cancer cells by activating expression of ephrin-B1. Cancer Sci 2018;109: 3159–70.

28. Ogi S, Fujita H, Kashihara M, Yamamoto C, Sonoda K, Okamoto I, Nakagawa K, Ohdo S, Tanaka Y, Kuwano M, Ono M. Sorting nexin 2-mediated membrane trafficking of c-Met contributes to sensitivity of molecular-targeted drugs. Cancer Sci 2013;104: 573–83.

29. Shtutman M, Baig M, Levina E, Hurteau G, Lim CU, Broude E, Nikiforov M, Harkins TT, Carmack CS, Ding Y, Wieland F, Buttyan R, et al. Tumor-specific silencing of COPZ2 gene encoding coatomer protein complex subunit zeta 2 renders tumor cells dependent on its paralogous gene COPZ1. Proc Natl Acad Sci U S A 2011;108: 12449–54.

30. Kim Y, Choi JW, Lee JH, Kim YS. MAD2 and CDC20 are upregulated in high-grade squamous intraepithelial lesions and squamous cell carcinomas of the uterine cervix. Int J Gynecol Pathol 2014;33: 517–23.

31. Zhang H, Zheng H, Mu W, He Z, Yang B, Ji Y, Hui L. DUSP16 ablation arrests the cell cycle and induces cellular senescence. FEBS J 2015;282: 4580–94.

32. Meng N, Glorieux C, Zhang Y, Liang L, Zeng P, Lu W, Huang P. Oncogenic K-ras Induces Mitochondrial OPA3 Expression to Promote Energy Metabolism in Pancreatic Cancer Cells. Cancers (Basel) 2019;12.

33. Zhao M, Li Y, Wei X, Zhang Q, Jia H, Quan S, Cao D, Wang L, Yang T, Zhao J, Pei M, Tian S, et al. Negative immune factors might predominate local tumor immune status and promote carcinogenesis in cervical carcinoma. Virol J 2017;14: 5.

34. Thomas G. Are we making progress in curing advanced cervical cancer? J Clin Oncol 2011;29: 1654–6.

35. Mizuno T, Suzuki N, Makino H, Furui T, Morii E, Aoki H, Kunisada T, Yano M, Kuji S, Hirashima Y, Arakawa A, Nishio S, et al. Cancer stem-like cells of ovarian clear cell carcinoma are enriched in the ALDH-high population associated with an accelerated scavenging system in reactive oxygen species. Gynecol Oncol 2015;137: 299–305.

36. Deonarain MP, Kousparou CA, Epenetos AA. Antibodies targeting cancer stem cells: a new paradigm in immunotherapy? MAbs 2009;1: 12–25.

37. Savage P. Chemotherapy curable malignancies and cancer stem cells: a biological review and hypothesis. BMC Cancer 2016;16: 906.

38. Fathy A, Abdelrahman AE. EZH2, Endothelin-1, and CD34 as Biomarkers of Aggressive Cervical Squamous Cell Carcinoma: An Immunohistochemical Study. Turk Patoloji Derg 2018;34: 150–57.

39. Sidney LE, Branch MJ, Dunphy SE, Dua HS, Hopkinson A. Concise review: evidence for CD34 as a common marker for diverse progenitors. Stem Cells 2014;32: 1380–9.

40. De Angelis ML, Francescangeli F, La Torre F, Zeuner A. Stem Cell Plasticity and Dormancy in the Development of Cancer Therapy Resistance. Front Oncol 2019;9: 626.

41. Steinbichler TB, Dudas J, Skvortsov S, Ganswindt U, Riechelmann H, Skvortsova, II. Therapy resistance mediated by cancer stem cells. Semin Cancer Biol 2018;53: 156–67.

42. Sharma SV, Lee DY, Li B, Quinlan MP, Takahashi F, Maheswaran S, McDermott U, Azizian N, Zou L, Fischbach MA, Wong KK, Brandstetter K, et al. A chromatin-mediated reversible drug-tolerant state in cancer cell subpopulations. Cell 2010;141: 69–80.

43. Talukdar S, Bhoopathi P, Emdad L, Das S, Sarkar D, Fisher PB. Dormancy and cancer stem cells: An enigma for cancer therapeutic targeting. Adv Cancer Res 2019;141: 43–84.

44. Zhang X, Meyerson, M. Illuminating the noncoding genome in cancer. Nature Cancer2020;1: 864–72.

45. Lawson DA, Kessenbrock K, Davis RT, Pervolarakis N, Werb Z. Tumour heterogeneity and metastasis at single-cell resolution. Nat Cell Biol 2018;20: 1349–60.

46. Tornesello ML, Faraonio R, Buonaguro L, Annunziata C, Starita N, Cerasuolo A, Pezzuto F, Tornesello AL, Buonaguro FM. The Role of microRNAs, Long Non-coding RNAs, and Circular RNAs in Cervical Cancer. Front Oncol 2020;10: 150.

47. Zhao G, Shi L, Qiu D, Hu H, Kao PN. NF45/ILF2 tissue expression, promoter analysis, and interleukin-2 transactivating function. Exp Cell Res 2005;305: 312–23.

48. Guan D, Altan-Bonnet N, Parrott AM, Arrigo CJ, Li Q, Khaleduzzaman M, Li H, Lee CG, Pe‘ery T, Mathews MB. Nuclear factor 45 (NF45) is a regulatory subunit of complexes with NF90/110 involved in mitotic control. Mol Cell Biol 2008;28: 4629–41.

49. Shamanna RA, Hoque M, Lewis-Antes A, Azzam EI, Lagunoff D, Pe‘ery T, Mathews MB. The NF90/NF45 complex participates in DNA break repair via nonhomologous end joining. Mol Cell Biol 2011;31: 4832–43.

50. Li N, Liu T, Li H, Zhang L, Chu L, Meng Q, Qiao Q, Han W, Zhang J, Guo M, Zhao J. ILF2 promotes anchorage independence through direct regulation of PTEN. Oncol Lett 2019;18: 1689–96.

## References

1. Jadaliha M, et al. A natural antisense lncRNA controls breast cancer progression by promoting tumor suppressor gene mRNA stability. PLoS Genet. 2018 Nov 29;14(11):e1007802.

2. Gao H, et al. Long noncoding RNA MAGI1-IT1 promoted invasion and metastasis of epithelial ovarian cancer via the miR-200a/ZEB axis. Cell Cycle. 2019 Jun;18(12):1393–1406.

3. Piipponen M, et al. p53-Regulated Long Noncoding RNA PRECSIT Promotes Progression of Cutaneous Squamous Cell Carcinoma via STAT3 Signaling. Am J Pathol. 2020 Feb;190(2):503–517. doi:10.1016/j.ajpath.2019.10.019. Epub 2019 Dec 12. Erratum in: Am J Pathol. 2020 Apr;190(4):916

4. Zhao K, et al. LncRNA FTX Contributes to the Progression of Colorectal Cancer Through Regulating miR-192-5p/EIF5A2 Axis. Onco Targets Ther. 2020 Mar 31;13:2677–2688.

5. Zhang F, et al. Long non-coding RNA FTX promotes gastric cancer progression by targeting miR-215. Eur Rev Med Pharmacol Sci. 2020 Mar;24(6):3037–3048.

6. Huo X, et al. FTX contributes to cell proliferation and migration in lung adenocarcinoma via targeting miR-335-5p/NUCB2 axis. Cancer Cell Int. 2020 Mar 23;20:89.

7. Wu C, et al. LINC00470 promotes tumour proliferation and invasion, and attenuates chemosensitivity through the LINC00470/miR-134/Myc/ABCC1 axis in glioma. J Cell Mol Med. 2020 Sep 11.

8. Huang W, et al. LncRNA LINC00470 promotes proliferation through association with NF45/NF90 complex in hepatocellular carcinoma. Hum Cell. 2020 Jan;33(1):131–139.

9. Yan J, et al. LncRNA LINC00470 promotes the degradation of PTEN mRNA to facilitate malignant behavior in gastric cancer cells. Biochem Biophys Res Commun. 2020 Jan 22;521(4):887–893

10. Gao H, et al. LncRNA LINC00974 Upregulates CDK6 to Promote Cell Cycle Progression in Gastric Carcinoma. Cancer Biother Radiopharm. 2019 Dec;34(10):666–670.

11. Tang J, et al. A novel biomarker Linc00974 interacting with KRT19 promotes proliferation and metastasis in hepatocellular carcinoma. Cell Death Dis. 2014 Dec 4;5(12):e1549.

12. Shi Y, et al. LINC00449 regulates the proliferation and invasion of acute monocytic leukemia and predicts favorable prognosis. J Cell Physiol. 2020 Oct;235(10):6536–6547.

13. Fang E, et al. Therapeutic Targeting of MZF1-AS1/PARP1/E2F1 Axis Inhibits Proline Synthesis and Neuroblastoma Progression. Adv Sci (Weinh). 2019 Aug 10;6(19):1900581.

14. Qian CJ, et al. LncRNA MAFG-AS1 Accelerates Cell Migration, Invasion and Aerobic Glycolysis of Esophageal Squamous Cell Carcinoma Cells via miR-765/PDX1 Axis. Cancer Manag Res. 2020 Aug 5;12:6895–6908.

15. Jiang B, et al. Up-regulation of miR-765 predicts a poor prognosis in patients with esophageal squamous cell carcinoma. Eur Rev Med Pharmacol Sci. 2018 Jun;22(12):3789–3794.

16. Wang X et al. A 15-lncRNA signature predicts survival and functions as a ceRNA in patients with colorectal cancer. Cancer Manag Res. 2018 Nov 16;10:5799–5806.

17. Gu X, et al. Construction and Comprehensive Analyses of a Competing Endogenous RNA Network in Tumor-Node-Metastasis Stage I Hepatocellular Carcinoma. Biomed Res Int. 2020 Feb 11;2020:5831064.

18. Koo KH, Kwon H. MicroRNA miR-4779 suppresses tumor growth by inducing apoptosis and cell cycle arrest through direct targeting of PAK2 and CCND3. Cell Death Dis. 2018 Jan 23;9(2):77.

19. Ye J, et al. Risk scoring based on expression of long non-coding RNAs can effectively predict survival in hepatocellular carcinoma patients with or without fibrosis. Oncol Rep. 2020 May;43(5):1451–1466.

